# Cell phone mobility data and manifold learning: Insights into population behavior during the COVID-19 pandemic

**DOI:** 10.1101/2020.10.31.20223776

**Authors:** Roman Levin, Dennis L. Chao, Edward A. Wenger, Joshua L. Proctor

**Affiliations:** Department of Applied Mathematics, University of Washington, Seattle, WA 98195, United States; Institute for Disease Modeling, Bill and Melinda Gates Foundation, WA 98109, United States

## Abstract

As COVID-19 cases resurge in the United States, understanding the complex interplay between human behavior, disease transmission, and non-pharmaceutical interventions during the pandemic could provide valuable insights to focus future public health efforts. Cell-phone mobility data offers a modern measurement instrument to investigate human mobility and behavior at an unprecedented scale. We investigate mobility data collected, aggregated, and anonymized by SafeGraph Inc. which measures how populations at the census-block-group geographic scale stayed at home in California, Georgia, Texas, and Washington since the beginning of the pandemic. Using manifold learning techniques, we find patterns of mobility behavior that align with stay-at-home orders, correlate with socioeconomic factors, cluster geographically, and reveal sub-populations that likely migrated out of urban areas. The analysis and approach provides policy makers a framework for interpreting mobility data and behavior to inform actions aimed at curbing the spread of COVID-19.

## 1 Introduction

The ongoing COVID-19 pandemic has had a devastating impact on mortality [1] and economic activity [2] leading to increased food insecurity, poverty, and gender inequity [3]. Most public health interventions attempting to arrest or mitigate the spread of the disease caused by the Severe Acute Respiratory Syndrome Coronavirus 2 are non-pharmaceutical interventions aimed at decreasing transmission by changing people’s behavior. For example, every state in the United States (US) issued mandatory or advisory *stay-at-home* orders between March and May of 2020 [4]. However, characterizing changes in behavior during the COVID-19 pandemic, whether due to adherence to stay-at-home orders, loss of employment, or non-pandemic-related factors, is challenging. In this article, we use cell-phone mobility data from SafeGraph Inc. to identify the heterogeneous mobility behaviors during COVID-19 in four states and reveal consistent motifs across states, within a state, and even within urban centers. Moreover, the modeling and analyses also point to geographic areas with populations that are young and highly mobile. We believe the approach and insights in the work could be leveraged by local public health officials to better target educational campaigns by geographic area and socioeconomic status.

Cell phone location data is a relatively new but promising way to quantify human movement. The locations of cell phones can be tracked by service providers or applications installed on the phones by users, but data that is shared with scientists is typically anonymized and aggregated to protect the privacy of individuals [5, 6]. Mobility data offers a unique measurement instrument to link public health statements and related legislative actions taken to reduce population mobility with an actual effect on population behavior. Cell-phone mobility data has provided early evidence that these orders were indeed associated with reductions in movement [7, 8, 9, 10, 11]; moreover, adherence was not uniform and may be associated with factors such as socioeconomic status and political leanings [7, 8, 9, 10, 11]. The keen interest in cell-phone mobility data to help inform policy makers during the COVID-19 pandemic has been widely discussed [12], with strong emphasis on the challenges facing data ascertainment bias, interpreting the link between mobility and behavior changes, and the lack of a single mathematical framework for analyzing this data [5, 6]. To date, most investigations of mobility data during the COVID-19 pandemic have compared summary statistics from mobility data, such as average cell-phone mobility within a region, between regions with different demographics. Here, we leverage the mobility data at full geographic and temporal resolution along with recently developed mathematical methods from dynamical systems and machine learning to identify patterns of behavior that are consistent across multiple geographic scales and provide insight into behavioral differences.

Analyzing and interpreting high-dimensional mobility time-series is a challenge. Model and dimensionality reduction has a rich history in the analysis of dynamical systems, with early theoretical work on bifurcation analyses enabling the categorization of qualitatively different dynamic regimes [13] to the more recent data-driven, equation-free approaches [14] enabled by advances in machine-learning and pattern analysis [15]. The standard approach typically involves a linear dimensionality reduction technique, such as the singular value decomposition (SVD) [16], in conjunction with a statistical clustering model to identify similarities across time series [17]. Despite the broad success of this approach, substantial limitations have been identified due to the underlying assumptions associated with the SVD and the mismatch with characteristics of data collected from a complex, temporally evolving system; this discrepancy has motivated the development of a diverse set of nonlinear dimensionality reduction techniques for time-series data [18]. Methods such as diffusion maps and Laplacian eigenmaps, popular in statistical and computational analyses [19, 20] and contained in a class of machine-learning methodologies called *manifold learning*, have been utilized by the dynamical systems community to identify nonlinear embeddings of the dynamics directly from observational data from the system [18]. Success has been demonstrated with these methods using data generated from simulation models [21]. Here, we leverage these methodologies to identify a lower-dimensional embedding of the mobility time-series data providing a framework that highlights common mobility behaviors at the census-block-group scale, identifies the geographic connectivity of behavior at different spatial scales, and reveals insights into epidemiologically relevant subpopulations.

The outline of this article is as follows: the following section of the paper provides a description of the SafeGraph mobility time-series and US census data (§2) along with the mathematical methods used for analysis §3. §4 highlights the heterogeneity and consistency of mobility patterns at the census block group level during the COVID-19 pandemic. In addition, we show these behaviors are correlated with socioeconomic indicators such as income and home ownership. We also describe a highly mobile population with distinctly different behavior revealed from the analysis. The final section §5 offers a discussion on how these findings provide insight into the connection between mobility, behavior, and transmission and how local public health officials can use this data to target information and education campaigns.

## 2 Data

### 2.1 SafeGraph mobility data

We obtained mobility data from SafeGraph, Inc. SafeGraph aggregates mobile device GPS data from various sources and produces anonymized datasets aggregated at the census block group (CBG) level. These data can be obtained free-of-charge for non-commercial use by joining their COVID-19 Data Consortium^1^. In this study, we estimate the number of people who stay at home each day by dividing the number of mobile devices that do not leave their homes by the total number of devices in each CBG (i.e., `completely home device count` divided by the `device count`) [22]. We used data covering 117 days of mobility, starting from February 23, 2020.

We define the daily proportion of devices seen near their homes to be the number of devices in each CBG detected in their home CBG (`destination cbg` = `origin census block group`) divided by the number of devices associated with the CBG (*device count*). The proportion of devices that are only detected *away from* their homes each day is 1 minus this proportion. Figure 1B. illustrates this daily stay-at-home fraction for five CBGs. We use the most recently released versions of the SafeGraph social distancing data, which is version 2.0 (“v2”) for dates before May 10, 2020 and version 2.1 (“v2.1”) for later dates [23]. Around May 17, SafeGraph began using “rolling windows” to assign the home census block group of devices instead of batch-updating only at the first of each month [24].

**Figure 1:**
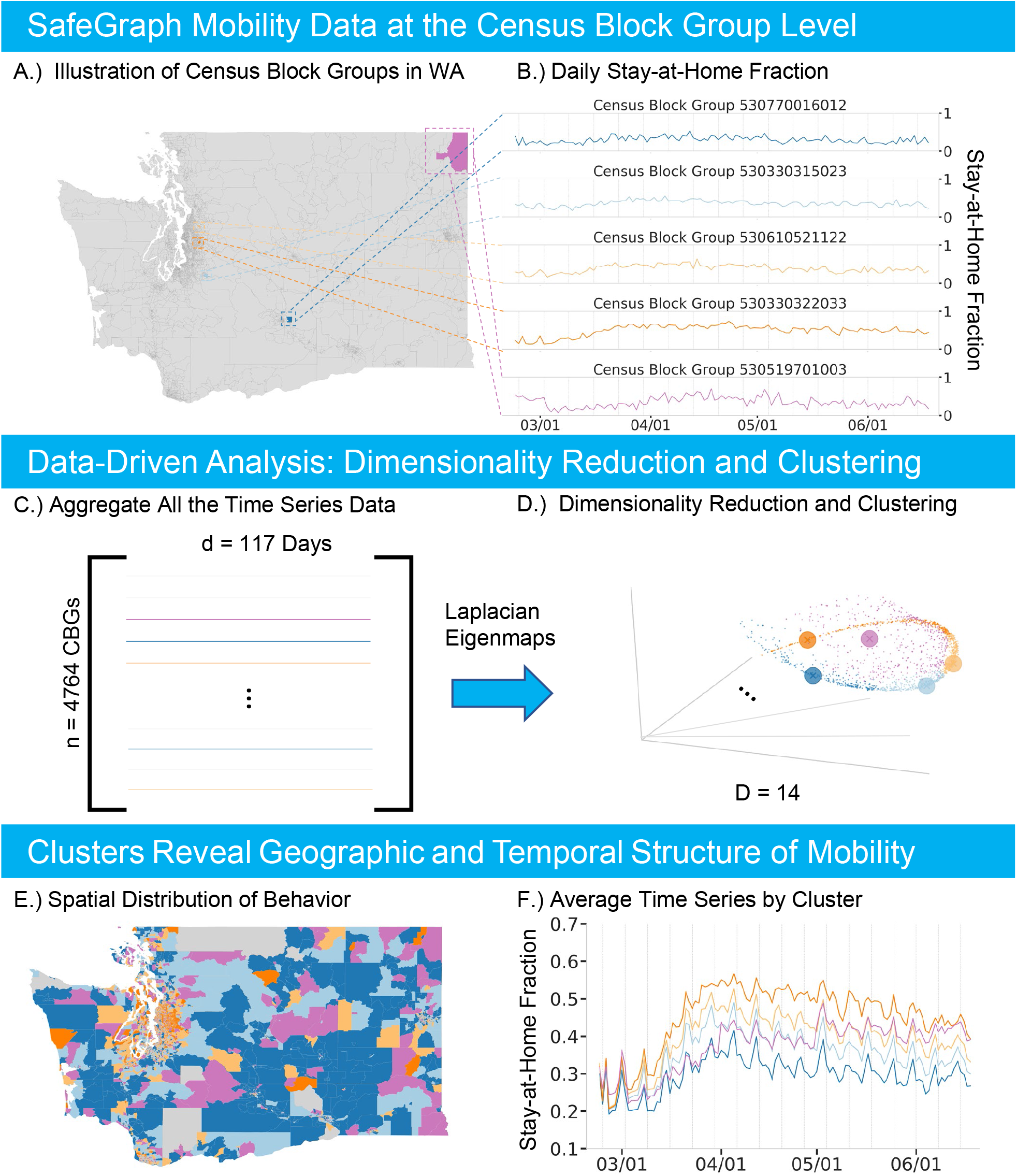
Data-driven analysis overview. A.) Example CBGs on the map of Washington. B.) Mobility time-series of the example CBGs. C.) Mobility time series are aggregated in a matrix form. D.) 3D visualization of the 14D Laplacian Eigenmaps embedding of the data with clusters highlighted in color. Large dots represent the example CBGs. E.) Clusters plotted on the Washington map. F.) Average mobility time-series for every cluster.

### 2.2 Census and geographic data

We obtained US population data from the 2018 American Community Survey (ACS) product of the US Census Bureau, accessed using the R package `tidycensus` [25]. We used Table B01001 for total population size and population by age estimates by CBG, Table B19013 for median household income, Table B14002 for number currently enrolled in college, Table B25008 for renter vs. owner-occupied housing units, and Tables B07201, B07202, and B07203 for “geographic mobility” (living in same house as last year). We computed a CBG’s population density by dividing the 2018 population estimate by the land area of the CBG as reported by the cartographic boundary files.

The US Census provides *cartographic boundary files*, which define simplified shapes of geographic entities designed for plotting. The 2019 shapefiles were downloaded from the US Census Bureau website^2^. The detailed map of Seattle was generated using ESRI’s “world topo map” [26] obtained using R’s `OpenStreetMap` package [27].

## 3 Methods

### 3.1 Linear dimensionality reduction: singular value decomposition

The singular value decomposition (SVD) is a standard linear matrix factorization technique that can be used to reduce the dimensionality of a data matrix [28, 16]. Using the SafeGraph time-series data (§2.1), we construct a mobility data matrix for each state. Each state’s data matrix has 117 columns (days of mobility data), but a different number of rows depending on the number of census block groups (Table 1). Figure 1C. illustrates the aggregation of mobility time-series into a data matrix for Washington state. For data matrix normalization, columns are mean subtracted. We perform a standard SVD to find a reduced order set of singular vectors and values for dimensionality reduction; see Supplement §1 for more details. The computational code to generate all results and figures in this article are publicly available [29].

**Table 1:**
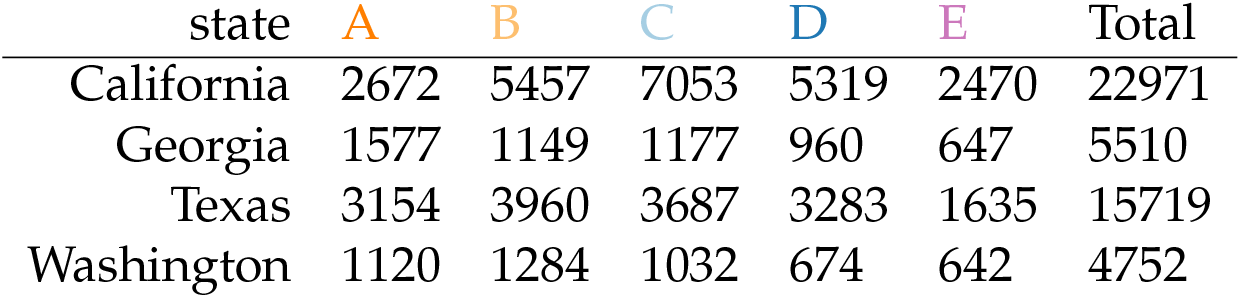
Number of census block groups (CBGs) in each cluster.

### 3.2 Manifold learning and nonlinear dimensionality reduction: Laplacian eigenmaps

Laplacian eigenmaps are a nonlinear manifold learning method that can identify a low-dimensional embedding which optimally preserves local structure of a high-dimensional data manifold [20]. To construct an *m*-dimensional embedding, the method uses *m* eigenvectors of the nearest-neighbors graph Laplacian corresponding to the smallest non-zero eigenvalues. The resulting embedding is optimal in the sense that “close” data points on the original manifold are represented by points that are close in the *m*-dimensional Euclidean embedding space; see equation (3.1) in [20] for more details. We also investigated a wide variety of other nonlinear dimensionality reduction techniques (Supplement §2). The Laplacian Eigenmaps algorithm was implemented using the `SpectralEmbedding` function from `sklearn.manifold` module of `scikit-learn` package [30] in `Python 3`. In this work, we used 50 neighbors for the `n neighbors` parameter. Varying the number of neighbors between 20 and 50 did not significantly change the Laplacian Eigenmap embedding for Washington state (Supplement §2).

The optimal effective dimensionality of the embedding was identified using the trustworthiness metric [31] which captures the extent to which a dimensionality reduction technique retains the local structure of the original data manifold from the higher-dimensional space. Trustworthiness was computed as a function of the Laplacian Eigenmap embedding dimensionality; a knee-point detection algorithm was then used to identify the optimal number of dimensions. The Supplement §2 provides a detailed description of this analysis for each state. To implement the trustworthiness metric, we used the function `trustworthiness` from `sklearn.manifold` of `scikit-learn` package [30] in `Python 3` with default parameters (5 neighbors, to capture the local structure). For the knee point detection, we used the Kneedle algorithm implemented in `kneed` package [32].

### 3.3 Gaussian Mixture Model Clustering

To interpret the low-dimensional structure revealed by Laplacian Eigenmaps, we apply Gaussian mixture model (GMM) clustering [33]. The GMM is a latent variable model which assumes that the data has sub-populations or clusters which follow Gaussian distributions with parameters governing the centroid location and covariance structure of each cluster. GMMs were implemented using the `mclust` [34] package of `R` (version 4.0 [35]). We leverage the probabilistic formulation of the GMM model as a natural way to quantify uncertainty of the cluster assignment. A more detailed description of the GMM model and uncertainty quantification is provided in Supplement §4. We used Bayesian Information Criterion (BIC) to identify the optimal number of GMM components [36, 37, 34]. We applied knee-point detection to the BIC curve using the Kneedle algorithm implemented in `kneed` package [32]; see Supplement §5 for more details.

### 3.4 Statistical Testing

To test the difference of the socio-economic covariates distributions between clusters, we used the Kolmogorov-Smirnov [38, 39] test as implemented in `kstest` function of `scipy.stats` package in `Python 3`. To determine the significance of trends of covariates associated with CBGs in clusters identified by the GMM, we used `jonckheere.test` from `clinfun` package [40] in `R` using 1000 permutations and assuming decreasing trends from cluster A to cluster D (Table 1).

## 4 Results

### 4.1 Stay-at-home levels and trends vary across CBGs, but there are distinct motifs that are consistent across four states

The SafeGraph stay-at-home data offers insight into the levels and trends of human mobility at the census block group (CBG) geographic scale during the 2020 COVID-19 pandemic in the United States (Figure 1). Nonlinear dimensionality reduction of the time-series data from Washington state revealed a low-dimensional embedding providing insight into the consistency of mobility behavior across CBGs (Figure 1D.). Moreover, the embedding and stay-at-home behaviors for Washington are qualitatively similar to those of Georgia, Texas, and California (Figure 2). The optimal embedding dimension was 14 for all four states, determined by the trustworthiness metric (§3.2, Supplement §2). A similar low-dimensional structure in the time-series data can be found with a diversity of nonlinear dimensionality reduction methods; see §4.5 and Supplement §2 for more details.

**Figure 2:**
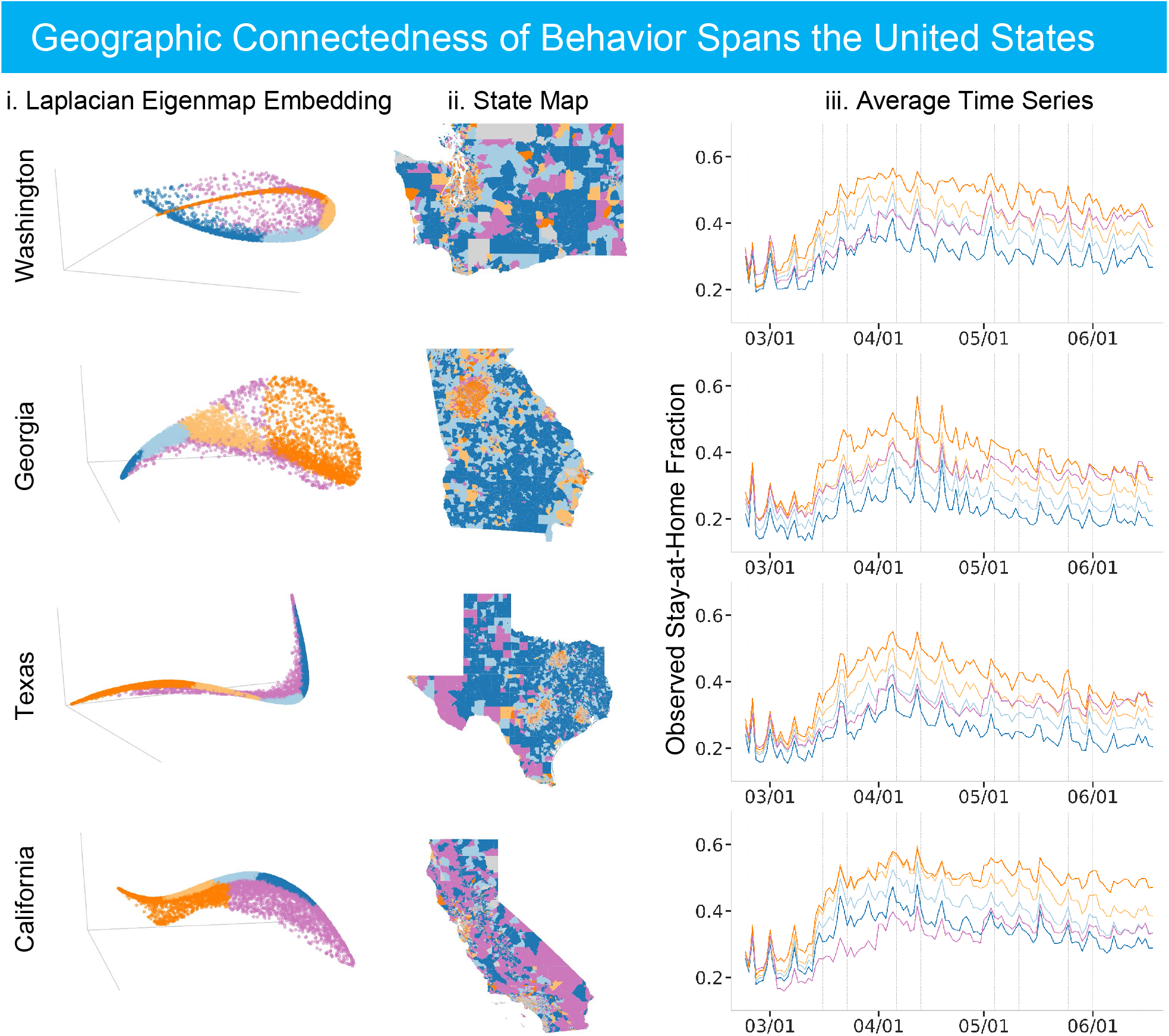
Results are consistent across four states. Column i. presents the 3D Laplacian Eigenmap visualization of the data manifold. Column ii. shows geographic maps. Column iii. presents average mobility time series for each cluster. Clusters are highlighted in color. It is noted that clusterig was done in 14D (optimal) embedding space.

The low-dimensional embedding provides insight into the similarity of stay-at-home behavior between CBGs. Figure 2 provides a visualization in three embedding dimensions of this coherent structure; note that for each state, certain CBG time-series are more similar to each other and the visualization indicates a large density of CBGs along a distinct, tubular data manifold. Fitting a Gaussian mixture model (GMM) to the stay-at-home time-series in the 14-dimensional embedding space identifies 4 clusters for California, Texas, and Washington and 5 clusters for Georgia (based on knee-point detection in BIC, see Supplement §5). For subsequent analyses, we chose 5 as the number of clusters for every state due to the optimal clustering for Georgia, comparisons across the four states (Table 1), and the geometric structure of the data (§4.2). Figure 2 illustrates how the clustering model groups CBGs in the embedding space (left column); the average mobility time-series for each cluster (right column) highlight the difference in stay-at-home behavior by cluster within a state and also the consistency across all four states. The cluster assignments were robust to model initialization (§4.5, Supplement §3) and had low associated uncertainty values (quantified in §4.5, Supplement §2, Supplement §4).

One clear difference between the clusters is their average level of mobility. For example in Washington, the average staying-at-home level increases from the CBGs in the dark blue cluster (cluster D) to the bright orange cluster (cluster A); see Figure 1F., representative CBG time-series in each cluster in Figure 1B., and Figure 2. The order of the clusters along the dense data manifold in the embedding space is aligned with their mean staying-at-home fraction (Figure 2). The average time series for clusters D through A do not intersect and are aligned in increasing order on the y-axis. However, the purple cluster E does not follow a similar trend with respect to the dense data manifold, nor the average time-series. For this cluster, we find that the fraction of the devices staying at home increases sharply in May 2020. A similar motif consistently occurs across each state (Figure 2); cluster E primarily captures outliers from the primary bulk trends that are continuously distributed across clusters A, B, C, and D. Those outliers are linked to a variety of important sub-populations, explored in more detail in §4.4.

The CBG clustering and average time-series by cluster also indicate that the change of behavior over time is different across clusters before April. The speed at which CBGs increased their stay-at-home behavior during a transition period between March and April (quantified by the slope of a linear fit of the CBG mobility time series during the transition period) is directly correlated with CBG cluster assignment (see Supplement §8). Moreover, the distributions of that speed are statistically significantly different: for every pair of clusters, we were able to reject the null hypothesis that the speed distributions were the same at the significance level *α* = 0.001 using Kolmogorov-Smirnov test. This is also directly evident by looking at this time period and the average stay-at-home trends by cluster (Figure 2). For example, the CBGs from the least mobile cluster A also increased their staying-at-home level the fastest.

### 4.2 CBGs with similar temporal mobility patterns are geographically consistent

The CBGs within each mobility cluster (defined in §4.1) are geographically connected and have consistent patterns across all four states. The second column of Figure 2 illustrates these broad trends which are most visually evident in the distinction between urban, peri-urban, and rural areas. For example in Washington, the Seattle area CBGs mostly belong to the bright orange and light orange most-staying-at-home clusters A and B, the same is true for nearby Bellevue and Redmond. Similarly, in Texas three large orange regions correspond to Dallas, Houston, and Austin. In Georgia, the distinct orange area on the map corresponds to Atlanta and in California we see orange colors around San Francisco, San Jose, and Los Angeles area. Likewise, blue colors – clusters C and D with lower stay-at-home levels – form continuous regions in rural areas on the state maps. CBGs that are close geographically tend to have similar mobility patterns.

Within each state, there is a stark contrast between urban and rural areas (Figure 2). For example in Washington, the large metropolitan areas around Seattle and Bellevue are colored orange (clusters A and B) as opposed to larger rural CBGs which belong to blue clusters (C and D). Large cities like Spokane or Yakima also have dense orange coloring (Figure 3) suggesting that changes in behavior within urban centers are similar despite being geographically quite distant from each other. The time series column of Figure 2 shows that urban areas (orange clusters A and B) stay at home significantly more than rural areas (blue clusters C and D). This observation is consistent across all four states.

**Figure 3:**
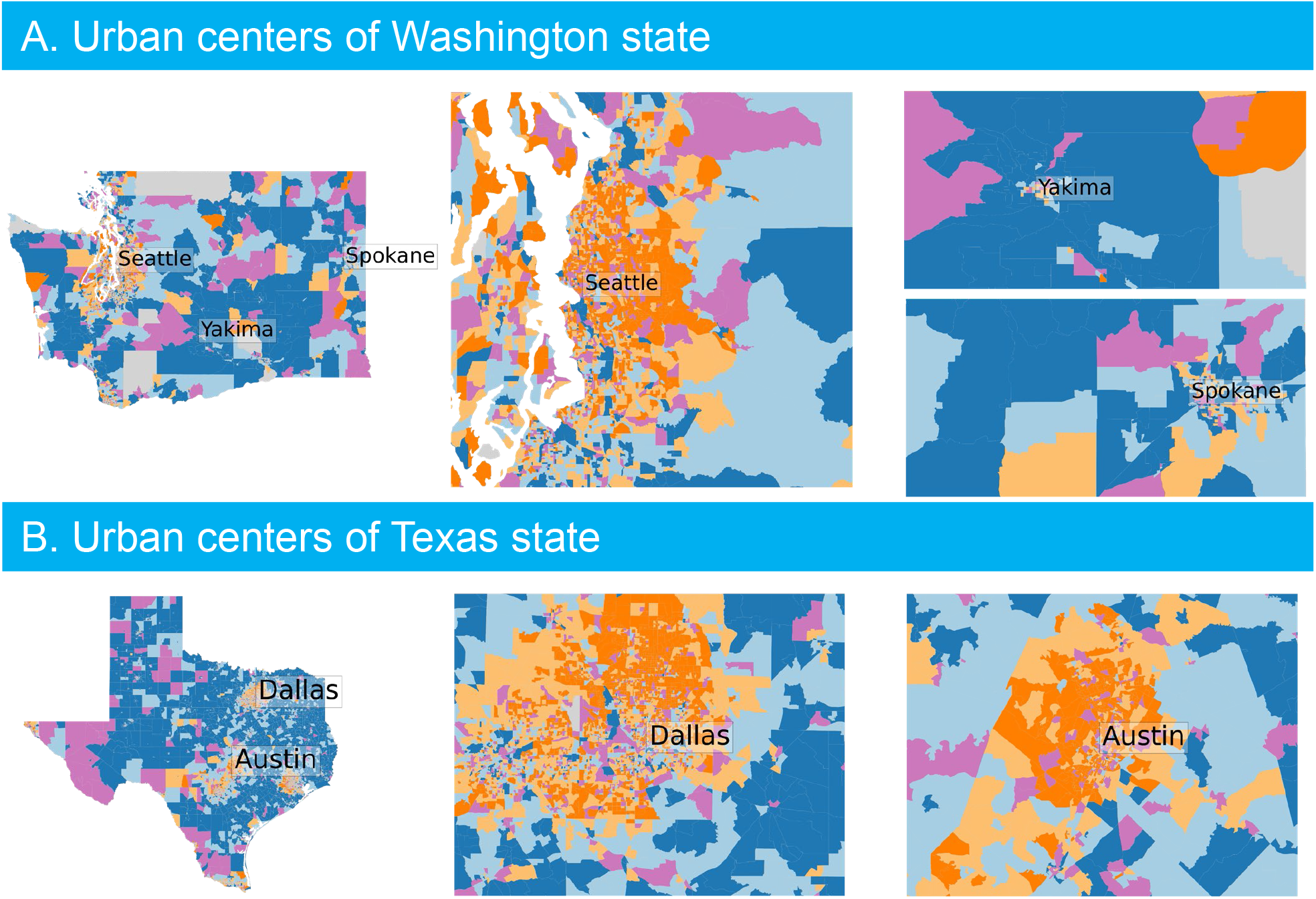
Clustering in metropolitan areas. Section A: clustering in Washington, Section B: clustering in Texas.

This analysis also identifies heterogeneity within the geographic scale of urban centers and rural areas. For example, in Dallas and Seattle there are urban CBGs that belong to blue clusters C and D indicating that they stay at home less than the surrounding areas (Figure 3). Moreover, populous cities such as Seattle, Atlanta, Austin and Dallas have distinct geographic groupings of CBGs for clusters A and B within the urban area (Figure 3, see Supplement §7 for clustering in Atlanta). The first column of Figure 2 clearly presents a smooth transition in the Laplacian Eigenmap embedding space between the bright orange cluster A that stays at home the most to the dark blue cluster D that stays at home the least. Remarkably, we observe the same on the geographic map. For example, there is a rough radial pattern around Dallas and Austin: bright orange CBGs densely cover the city center and are replaced by light orange, then light blue and eventually dark blue as the distance from the city center increases (see Figure 3). That is, the transition is quite consistent – it covers the intermediate colors and the stay-at-home level gradually decreases as distance from the city increases suggesting a more nuanced interpretation about the continuity of behavior across CBGs within urban centers supported by the geometric structure of the data manifold. In the greater Seattle area, the transition is substantially less pronounced especially moving eastward from downtown; note that both a large urban area (Bellevue) and suburb (Redmond, the home to Microsoft) exist to the east of Seattle, both with a higher income population.

Despite the optimal number of clusters being 4 or 5 for each state (§4.1), relaxing this criteria and allowing for more clusters provides more granular information within and around urban areas while maintaining consistency with the optimal GMM. This also follows the intuition provided by the illustrations of the nonlinear embedding in three dimensions (left column, Figure 2); namely, the embedding is broken into finer grained clusters enabling higher resolution comparisons between CBGs. Supplement §6 provides details on altering the number of clusters. Further, a continuous mapping of the data along the dense tubular structure of the data manifold shows the smooth transition across urban, peri-urban, suburban, and rural areas; see Supplement §6 for more details. In contrast, the purple cluster E is not wholly on the tubular structure and does not exhibit the same geographically connected characteristics as the other clusters. More detail is provided on this cluster and the possible difference in its subpopulation structure in §4.4.

### 4.3 Income, population density, and behavioral data are correlated

Clusters A and B, which on average stayed home the most, included the most densely populated CBGs, while clusters C and D included the more sparsely populated ones (Figure 4). High population density is generally an indication of urban populations and low density of rural areas (see maps in Figure 2). The CBGs in clusters A and B also had the highest median household incomes (Figure 4). In all states, the median stay-at-home fraction, population density, and household income of CBGs had a consistently decreasing trend from clusters A to D, and the Jonckheere–Terpstra test rejects the null hypothesis that these four clusters come from the same distribution of values (p*<* 0.01). Cluster E did not follow these trends and appeared to cover a wider range of values (Figure 4).

**Figure 4:**
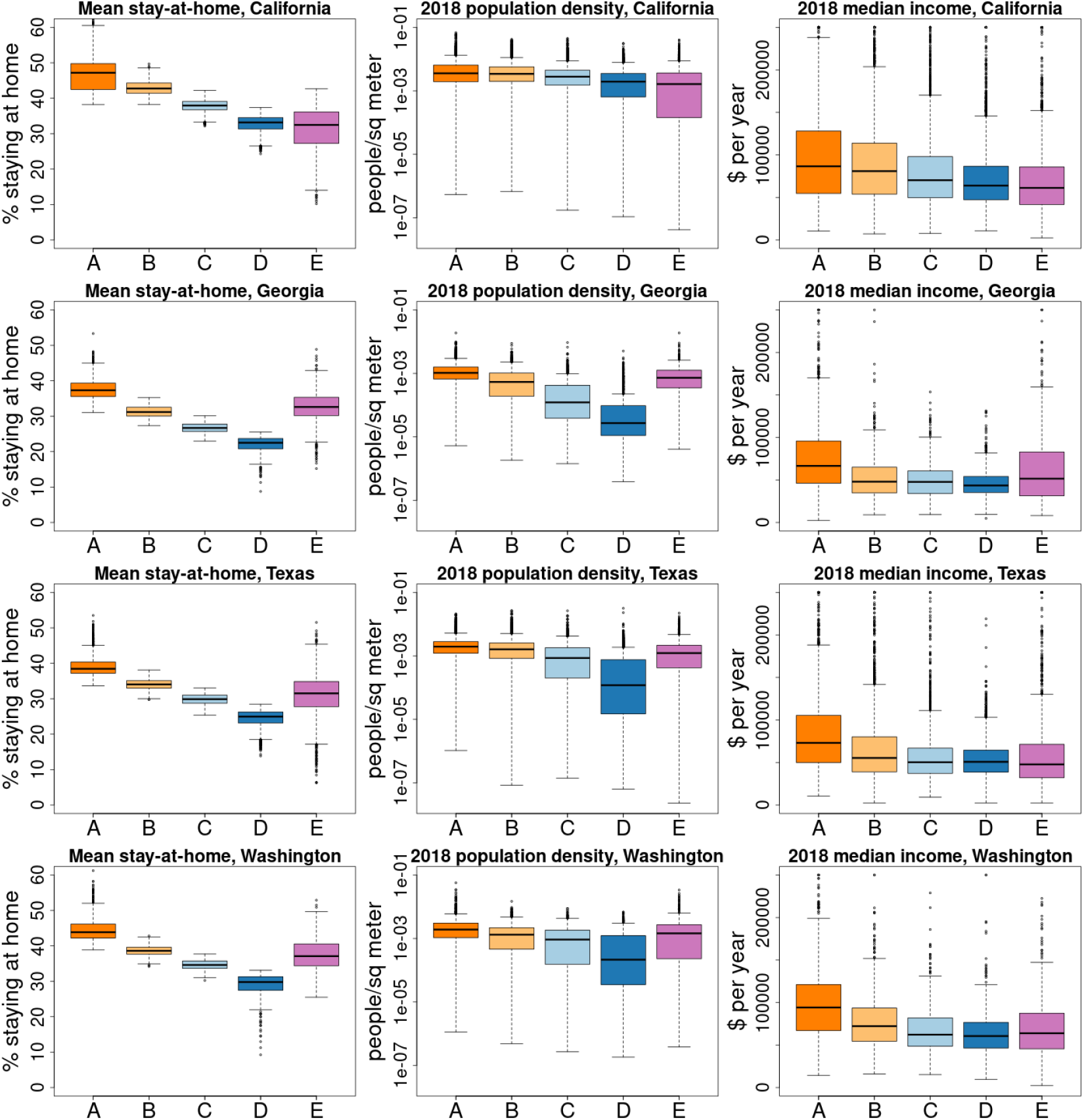
Distribution of stay-at-home behavior and demographic covariates by cluster. The boxplots present the interquartile range (boxes) and median values (center horizontal lines) of the covariate values for CBGs in each of the five clusters. Whiskers span the 95% range. The “mean stay at home” fraction of a CBG is the mean of the daily percent of mobile devices that stayed completely at home during the time period analyzed.

Cluster E has a higher proportion of people who we expect to have high “geographic mobility” (i.e., change residences frequently). Using 2018 ACS estimates, CBGs with a low proportion living in the same house in the previous year or a high proportion of renters, people enrolled in undergraduate or professional degree programs, or who are “young adults” (18 to 29 years old) tended to be in cluster E (Figure 5). In California, the proportion of people with high geographic mobility appears to be higher in cluster A than in cluster B.

**Figure 5:**
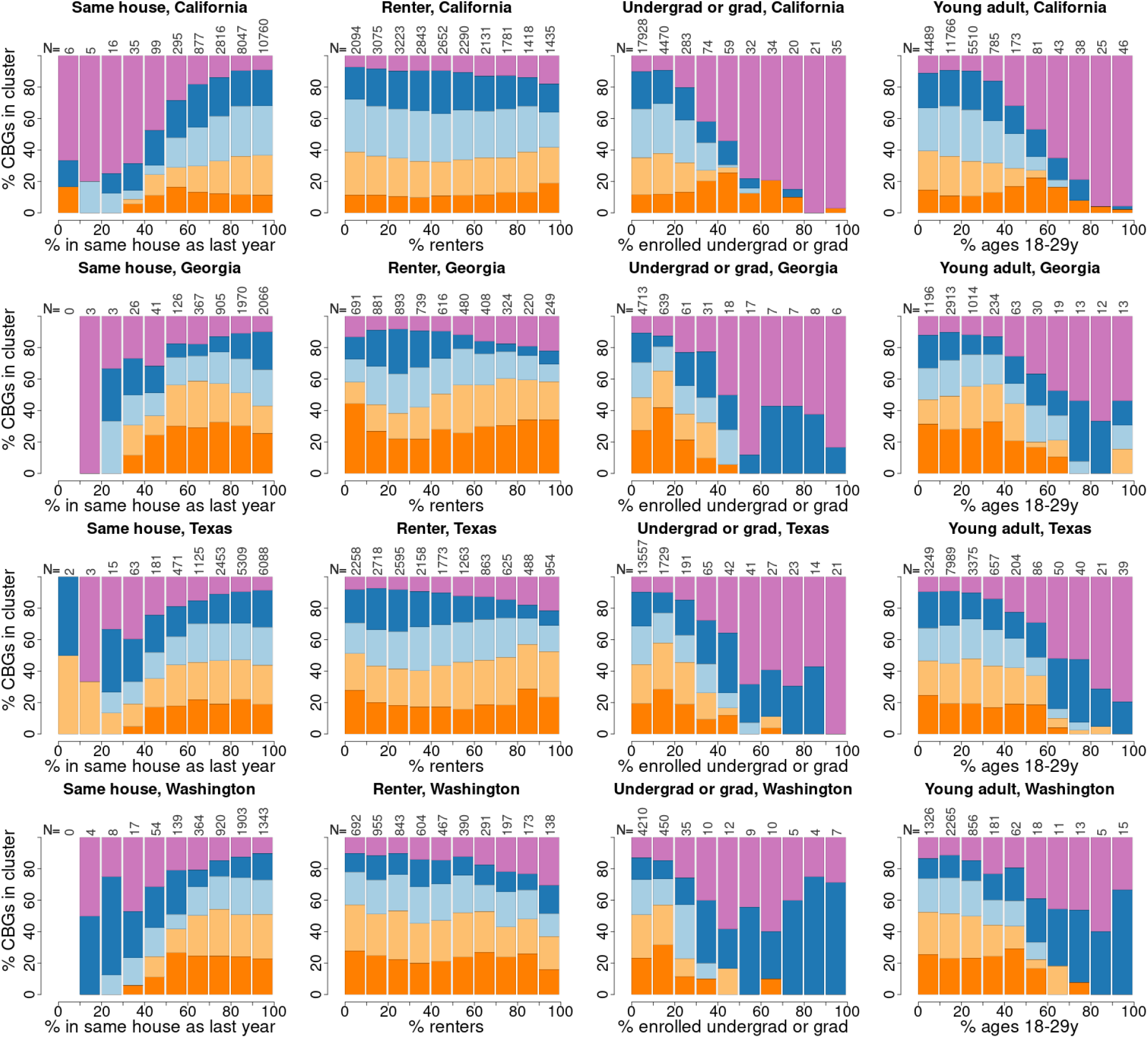
Proportions of census block groups in each cluster by population characteristics associated with geographic mobility. The fraction of a CBG’s population associated with the characteristic is plotted on the x-axis. CBGs are partitioned into 10 equally-spaced bins, defined by the proportion of each CBG’s population having the characteristic in 10% increments. The numbers of CBGs belonging to each bin are printed along the top of each panel. The proportion of CBGs in each cluster is plotted as vertically stacked bars for each bin (with cluster A in dark orange on the bottom through cluster E in purple on top).

Upon closer investigation of the location of clusters in the city of Seattle, Washington, the spatial distribution of clusters D and E is consistent with the associations described above (Figure 6). The area surrounding the University of Washington, where a large number of undergraduate and graduate students live, is in cluster E, while the University itself is in cluster D (Figure 6, center of map). Cluster E also includes downtown and Lake Union, where a recent influx of young tech workers fueled the development of new apartments. Interestingly, in addition to students and young tech workers in Seattle, cluster E also indicates some very high median income populations on the waterfront of Bellevue and Kirkland that were also highly mobile during this period. Cluster D includes “SODO”, the industrial area southwest of downtown, which is less affluent than the populations to the west, east, and north.

**Figure 6:**
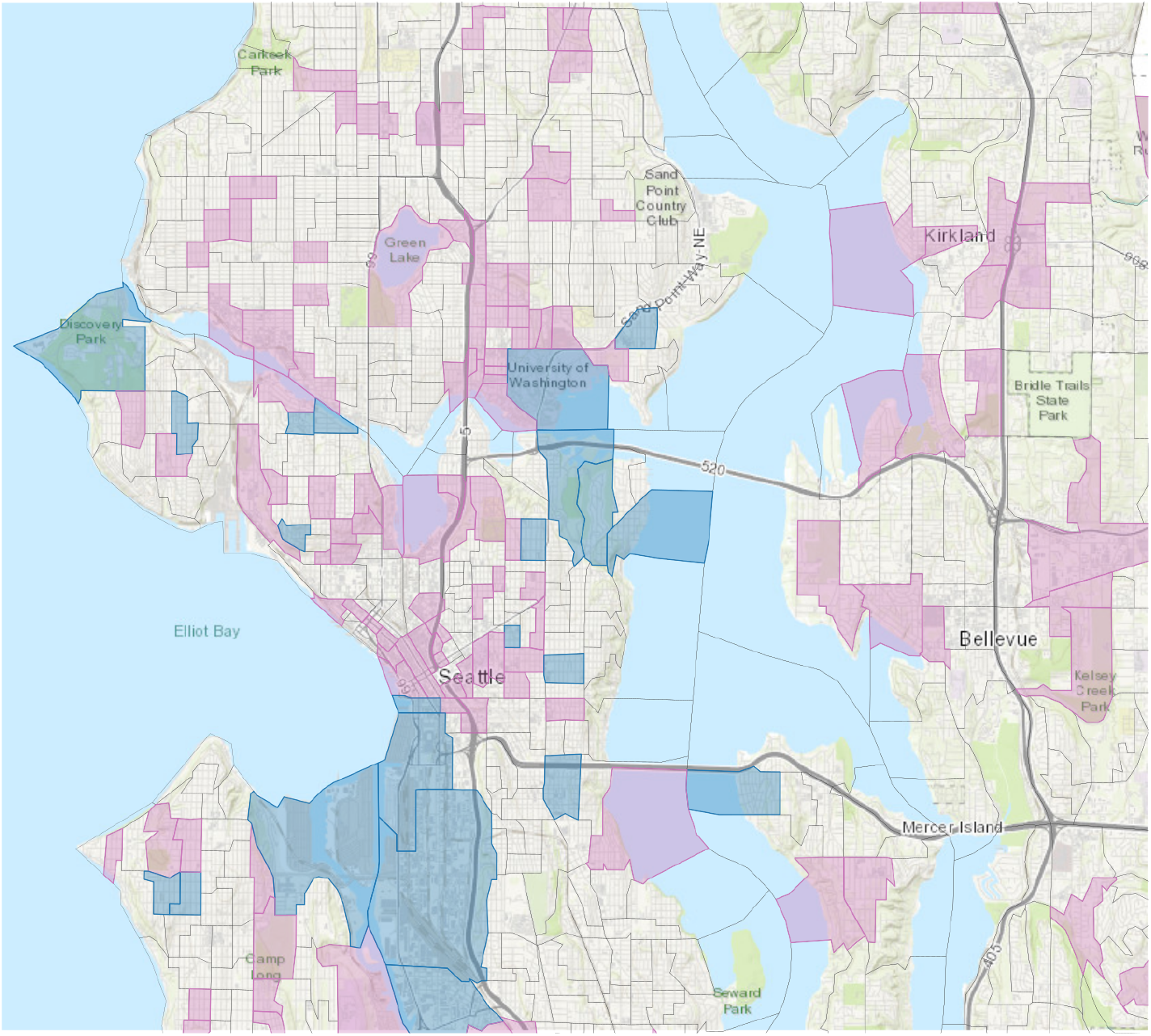
Clusters in the Seattle metropolitan area. Census block group boundaries are outlined. CBGs belonging to clusters D and E are highlighted in dark blue and purple, respectively.

### 4.4 Analysis reveals areas with likely population turnover early in the pandemic

The available SafeGraph dataset does not allow one to track the movements of individuals, but there are trends consistent with high population turnover. One can track the number of mobile devices that are detected by SafeGraph each day but not in their “home” CBG on a given day, which we call “never-near-home” devices. These devices could be on a trip away from home or they could have moved away entirely.

In March, the fraction of never-near-home devices was highest in cluster E (Figure 7 and Supplement §8). On April 1 and again on May 1, the number of devices never near home drops sharply in cluster E but not in the other clusters. This behavior is consistent with the owners of these devices moving to a new residence and SafeGraph re-assigning these devices to the new residence on the first day of a subsequent month. SafeGraph defines a person’s “home” to be the location where the mobile device is detected most at night (from 6pm to 7am) over a 6-week period [23]. If a person spends enough time in a new location, that new location can become the device’s “home”. These home locations were updated by SafeGraph at the start of each month until mid-May, when SafeGraph changed its procedure for assigning home locations to devices [24, 41]. The high proportion who were never near their “homes” in March and April and the sharp drops in these fractions on April 1 and May 1 in cluster E, and to a lesser extent in cluster D, are consistent with this population moving away. In California, cluster A also has a noticeable decline on May 1 (Supplement §8), which could indicate a high-income group that is geographically mobile. If a large number of people in a region move away, the devices will appear to be “away from home” because their home locations are out-of-date. These clusters will appear to be staying at home less than they really are. This batching artifact appears to be resolved in May 2020, and the stay-at-home fraction in cluster E rises relative to the other clusters.

**Figure 7:**
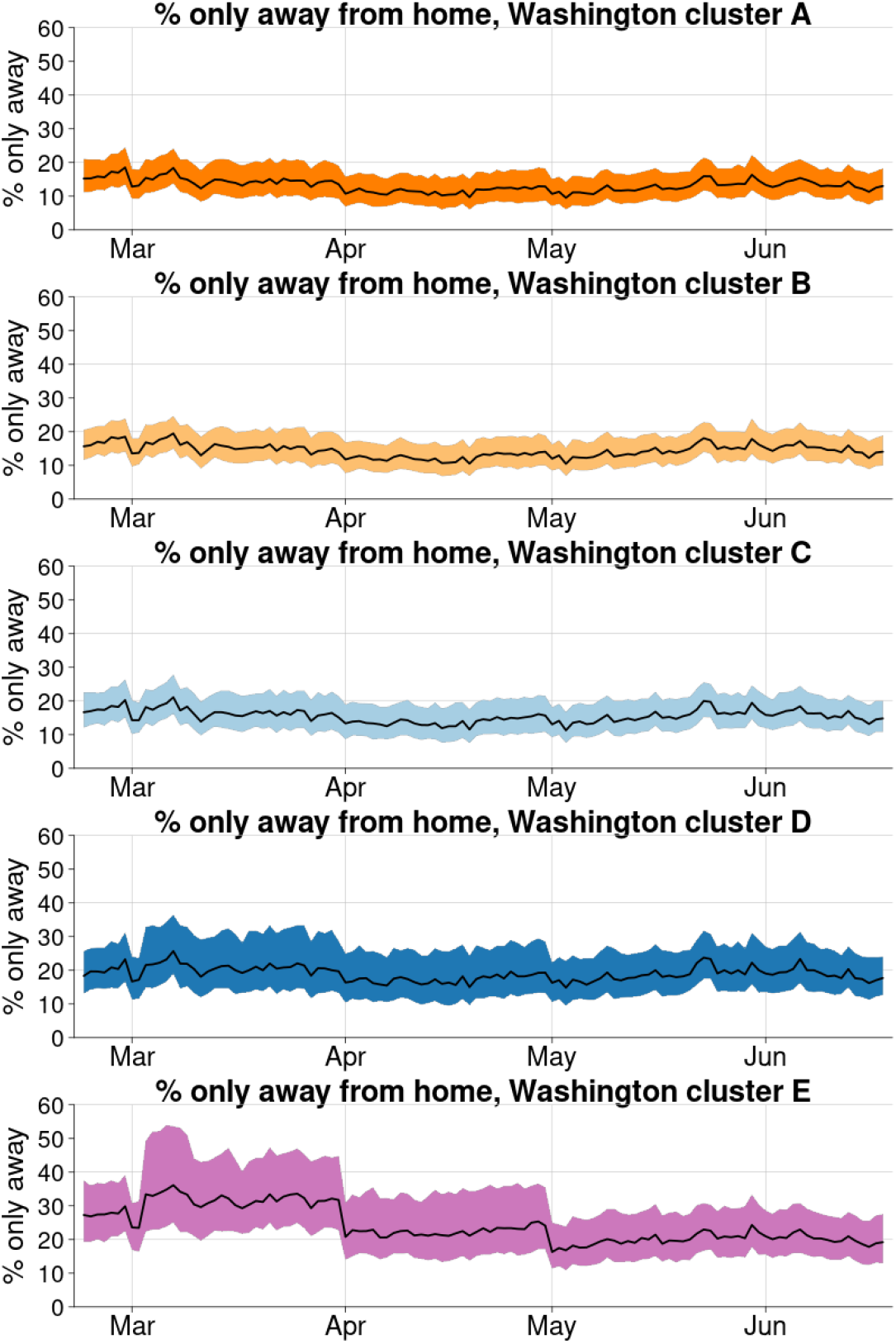
The fraction of devices that are *only* away from their homes each day. The medians and inter-quartile range of each day’s values are shown for each cluster in Washington State.

### 4.5 The nonlinear embedding and clustering results are robust

We investigated the sensitivity of our results to the methodological approach. The cluster assignment for the GMM in the 14 dimensional embedding space is robust. For every state, the maximum uncertainty is below 50% while the third quartile of the cluster assignment uncertainty is close to zero; at least 75% of the CBGs are well separated by a GMM in the 14-dimensional embedding space (see Supplement §2, Supplement §3). However, given the intrinsic structure of the data (Figure 2), the quantification of uncertainty for clustering is consistent with the geometry of the mobility data being more continuous than discrete across clusters A,B,C,D. For example, the uncertain CBG assignments are linked to the boundaries of clusters on or near the tubular data structure. The number of clusters and cluster assignments were optimized according to a standard approach which balances model fit and parsimony(§3.3), but the number of clusters could be changed depending on a desired level of granularity or continuous colormap could be used (Supplement §6). It is worth noting in contrast that clustering in the linearly reduced space is highly uncertain; for more details see Supplement §1.

We also found consistent results using alternative dimensionality reduction techniques such as Locally Linear Embedding and Isomap. For both Isomap and Locally Linear Embedding, a similar dense tubular data manifold was present in the lower dimensional embedding space (Supplement §2). Furthermore, the trustworthiness metric and knee point detection indicated that all three manifold learning methods agreed that the effective dimensionality of the embedding was between 14 and 16 (Supplement §2).

## 5 Discussion

Our results are consistent with other studies linking demographic characteristics to cellphone mobility data during the 2020 SARS-CoV-2 pandemic. Two recent studies using data from SafeGraph found mobile devices from areas with higher median household incomes stayed home more than devices from lower-income areas [9, 11], and this trend occurs in other mobile device datasets [7]. These studies hypothesize that the relationship between income and mobility is due in part to the ability of people with high-paying jobs to work from home. A survey found that about half of adults in Seattle switched to telework because of COVID, with high-income households making the change far more than lower-income (79.3% in households making *>*$150,000 per year and 23.5% among those making *<*$50,000) [42]. A recent study using another source of cell phone mobility data found that mobility was reduced more in urban than rural England [10], indicating that these trends could generalize beyond the United States.

Several related studies cluster mobility time series by a single demographic characteristic selected *a priori*, such as income [7, 9, 11] or population density [10] or party affiliation [8], to demonstrate behavioral differences with respect to that characteristic. Alternatively, one could reduce the time series to a summary statistic, such as average stay-at-home level over a particular time window, and study the relationship between that metric and several demographic covariates. In contrast, our methodological approach is broader; we measure similarities between complete time series, which allowed us to identify population clusters that had a distinct *change* in behavior, which would have been hidden if we had clustered by average behavior over time. Notably, we have identified features in the SafeGraph stay-at-home data that strongly suggests a mass migration out of several major metropolitan areas, especially in CBGs that have high proportions of young adults, renters, or students (§4.4). The closure of college of campuses and widespread job losses in March and April led many, especially young adults, to move [43, 44]. Moreover, the map presented in Figure 6 also matches our own intuition of where students of the University of Washington live, both adjacent to the university as well as more distant rental housing along bike and metro commuting lines (all authors of this manuscript live in the greater Seattle area). Similarly, the high-migration census block groups identified near South Lake Union tends toward a younger, professional population working at technology companies such as Amazon, and CBGs on the waterfront with high income populations in nearby cities such as Bellevue and Kirkland have a similar outward migration trend.

Identifying the population that moved early in the pandemic is a direct consequence of using a data-driven, equation-free approach. The approach has been integral to revealing the heterogeneity, but also the consistency, of mobility patterns across California, Georgia, Texas, and California; it has enabled a multi-scale geographic perspective on behavior allowing insights at the state, urban-rural, peri-urban, and suburban scale. Recent efforts have also utilized clustering of mobility time-series data specifically for analyzing SafeGraph stay-at-home data in Atlanta [9]. Our approach, though, is substantially broader in scope; identifying the low-dimensional embedding of the data enables a characterization of the geometric structure and the relatedness of each CBG mobility behavior. Moreover, we found utilizing a nonlinear dimensionality reduction techniques such as Laplacian Eigenmaps for analyzing mobility time-series data is essential (Supplement §1) mirroring recent developments from dynamical systems focused on the development of equation-free methods for analyzing measurement data collected from complex systems [18]. We have also leveraged clustering as a tool to interpret the similarity of mobility behavior between CBGs even in the reduced nonlinear embedding; we found that clusters allowed for comparisons of mobility characteristics (Figure 1), generalization across four states (Figure 2), and also correlation with socioeconomic factors (Figure 4 and Figure 5). The nonlinear embedding, however, offers a more nuanced perspective about the similarity of mobility behavior between CBGs. For example, the visualization in three dimensions and the clustering results suggests a much smoother and continuous geometric structure of relatedness for CBGs assigned to clusters A,B,C,D (Figure 2 and Supplement §6). This helps frame the clustering results and socioeconomic factor correlation analysis. Further, the embedding provides a richer characterization of the underlying complexity in mobility behavior.

We acknowledge several limitations of the mobility data and challenges in linking behavior to demographic variables. SafeGraph aggregates mobility data from many uncoordinated sources on the locations of millions of cell phones. These phones are not systematically tracked, and the GPS data might not be precise. The data are then aggregated by census block group and filtered to preserve the privacy of the mobile device owners. It is difficult to ascertain how well a set of mobility data represents the general population [5, 6]. Different states, and segments of the population, have different levels of coverage that are hard to correct for [45]. This is further complicated by likely gaps in coverage for high-risk populations such as migrant agricultural workers. However, the associations we found between mobility and other factors are consistent with those found in other datasets and are quite plausible. We studied the fraction of mobile devices that stayed at home each day, but this is just one metric than can be derived from the mobility data. Other measures, such as the mean length of time spent outside the home, the distance traveled from the home, or even the number of trips to stores, could provide additional insight into the population’s response to the pandemic. The demographic data in this study was from the 2018 American Community Survey, which we believe generally reflects the population in 2020 but might not accurately characterize the demographics of the most rapidly changing areas. We cannot establish the direct cause of the differential reductions in mobility using these data. We use demographic and socioeconomic variables at the census block group level, which could lead us to ecological fallacies, and many of these variables are tightly linked, thus, disentangling their effects is not straightforward and could be counterproductive.

Despite these challenges, population mobility data and connections to behavior can inform public health policy makers. Population behavior is a key component to understanding disease transmission dynamics; mobility data and the methods contained in this article helps quantify the change in population behavior during the pandemic. Policy makers can use this tool to assess the impacts of policy, especially important as COVID-19 cases start to resurge in the United States during a period of quarantine fatigue. We have also demonstrated that these data, analyses, and setting-specific information can provide epidemiologically relevant insights such as we uncovered around urban migration events. We believe the research in the article will provide insights for policymakers as they consider more modern, optimized, and targeted intervention strategies.

## Data Availability

All data used in this article are publicly available. We obtained mobility data from SafeGraph, Inc. SafeGraph aggregates mobile device GPS data from various sources and produces anonymized datasets aggregated at the census block group (CBG) level. These data can be obtained free-of-charge for non-commercial use by joining their
COVID-19 Data Consortium. We obtained US population data from the 2018 American Community Survey (ACS) product of the US Census Bureau, accessed using the R package tidycensus. The US Census provides cartographic boundary files, which define simplified shapes of geographic entities designed for plotting.

https://www.safegraph.com/covid-19-data-consortium

https://CRAN.R-project.org/package=tidycensus

## Author contributions

DLC, EAW, and JLP conceived the study, DLC and RL conducted the analyses, DLC, RL, EAW, and JLP wrote the manuscript, DLC, RL, and JLP wrote the supporting material.

## Acknowledgements

The authors would like to thank Dr. Marita Zimmermann, Dr. Roy Burstein, and Dr. Mike Famulare for helpful conversations around COVID-19 case data, correlations with socioeconomic data, and cell phone data.

## Footnotes

1 https://www.safegraph.com/covid-19-data-consortium

2 https://www.census.gov/geographies/mapping-files/time-series/geo/cartographic-boundary.html

## Notes

### Competing Interest Statement

The authors have declared no competing interest.

### Funding Statement

No external funding was received for this work.

