## Supplement for "Cell phone mobility data and manifold learning: Insights into population behavior during the COVID-19 pandemic"

### 1 Linear dimensionality reduction and clustering

We used the singular value decomposition (SVD), closely related to principal component analysis, as a linear method to reduce the dimensionality of the mobility data matrix. Figure S1 illustrates that more than 60 singular vectors (principal components) are required to reach 90% of the explained variance. This singular value distribution suggests a marginal benefit from linear dimensionality reduction. Moreover, applying a GMM to the data in the transformed space, we obtained highly uncertain GMM cluster assignments (see Table 1 and Supplement 4 for uncertainty quantification). Figure S1 provides a 3D visualization of clustering in the first three singular vectors. This provides an illustration for why cluster assignment is uncertain and changes depending on the model initialization. Linear dimensionality reduction and GMM clustering is not an effective method for the mobility dataset motivating the use of manifold learning methods.

| 0% | 25% | 50% | 75% | 100% |
|---|---|---|---|---|
| 3.38e-08 | 1.91e-01 | 3.53e-01 | 4.97e-01 | 7.46e-01 |

Table 1: Quartiles of the cluster assignment uncertainty based on a GMM with 11 clusters and the SVD with 8 modes.

### 2 Nonlinear Dimensionality Reduction Methods

We investigated a variety of nonlinear dimensionality reduction methods: t-SNE, locally linear embedding (standard, modified, and Hessian), Isomap, Spectral Embedding, local Tangent Space alignment, and multi-dimensional scaling. We found that Laplacian Eigenmaps [1], Locally Linear Embedding [5], and Isomaps [8] reduced the dimensionality of the data and identified a consistent tubular dense structure in the data (Figure S2). Figure S2 presents 3D representations of low-dimensional embeddings for each of the methods on the example of Washington State.

As described in Methods of the main manuscript, we chose the optimal embedding dimensionality based on the knee-point of the trustworthiness metric as a function of number of dimensions. All three methods qualitatively agree on the intrinsic dimensionality of the mobility data with the optimal dimensionality identified to be between 14 and 18 dimensions (see Figure S3 for the trustworthiness metric plots for Washington State). All three methods capture the structure of the data well and produce cluster assignments with significantly lower associated uncertainty than linear dimensionality reduction (see Table 2).

| Method | 25% | 50% | 75% | 100% |
|---|---|---|---|---|
| Laplacian Eigenmaps | 0 | 0 | 1.11e-15 | 4.99e-01 |
| Locally Linear Embedding | 1.65e-06 | 6.16e-04 | 4.30e-02 | 6.18e-01 |
| Isomap | 4.79e-08 | 5.90e-05 | 4.32e-03 | 4.97e-01 |

Table 2: **Quartiles of uncertainty of the cluster assignment based on GMM with 5 clusters.**

We selected Laplacian Eigenmaps as the primary methodology because it produced the least uncertain GMM cluster assignment and the clustering was robust to perturbations in the method's single hyperparameter, `n_neighbors` (Figure S4). Trustworthiness was computed as a function of the Laplacian Eigenmap embedding dimensionality; a knee-point detection algorithm was then used to identify the optimal number of dimensions. Figure S5 shows the optimal Laplacian eigenmaps dimensionality is 14 for every state. Note that California is different in that there are two possible knee points: one that is consistent with other states at 14 and another at 44 dimensions. We compared clustering results for these two knee points (Figure S6) and identify that the cluster assignments are similar. Therefore, in our main analysis we used 14D Laplacian eigenmap embedding for California.

### 3 Robustness of GMM fitting

The underlying objective function for the standard implementation of a Gaussian mixture model is not convex. We ensured that the GMM clustering produced consistent and robust results by re-initializing the fitting algorithm many times. As an example, Figure S7 illustrates the GMM fit for six different initializations. We observe that the results are qualitatively similar with the main difference that the sparse region sometimes is identified as a separate cluster or divided into two clusters.

### 4 GMM Model and Uncertainty Quantification

Gaussian mixture model (Section 11.2 in [4]) is a latent variable model which assumes that the data has $K$ sub-populations which follow Gaussian distributions with parameters $\mu_k, \Sigma_k$ respectively and that a latent discrete variable $z_i \in \{1, \dots, K\}$ controls which sub-population a data point $x_i$ comes from. If $\pi$ corresponds to the probability mass function of $z_i$, then the GMM model has the form:

$$p(x_i|\theta) = \sum_{k=1}^{K} \pi_k \mathcal{N}(x_i|\mu_k, \Sigma_k),$$

where $\theta$ stands for the set of all parameters of the model and $\mathcal{N}(x_i|\mu_k, \Sigma_k)$ is the probability density function of normal distribution. We note that GMM could be seen as a generalization of the famous K-means clustering algorithm [3].

The probabilistic formulation of the GMM model provides a natural way to quantify uncertainty of the cluster assignment. Using Bayes' Theorem, the posterior probability $p(z_i = k|x_i, \theta)$ that point $x_i$ belongs to cluster $k$ can be computed as follows:

$$p(z_i = k|x_i, \theta) = \frac{p(z_i = k|\theta)p(x_i|z_i = k, \theta)}{\sum_{k'=1}^{K} p(z_i = k'|\theta)p(x_i|z_i = k', \theta)} = \frac{\pi_k \mathcal{N}(x_i|\mu_k, \Sigma_k)}{\sum_{k'=1}^{K} \pi_{k'} \mathcal{N}(x_i|\mu'_k, \Sigma'_k)}.$$

Then, the amount of uncertainty $\epsilon_i$ in the cluster assignment of point $x_i$ could be computed as

$$\epsilon_i = 1 - \max_k p(z_i = k|x_i, \theta).$$

We note that the above formula assumes that the cluster assignment is computed as

$$z_i^* = \arg\max_k p(z_i = k|x_i, \theta).$$

### 5 GMM Model Selection

We used Bayesian Information Criterion (BIC) to identify the optimal number of GMM components [6, 2, 7]. BIC is based on a penalized form of the log-likelihood. As the likelihood increases with the addition of more components, a penalty term for the number of estimated parameters is subtracted from the log-likelihood [7]. To find the optimal number of GMM components, we applied knee-point detection to the BIC curve (Figure S8). Note that in the `mclust` implementation, higher BIC values correspond to better models. The optimal number of clusters turned out to be 4 for Washington, Texas, and California and 5 for Georgia. We decided to use 5 clusters for every state in the main manuscript for consistency across the states and to leverage optimal results for Georgia.

### 6 Altering the Number of Clusters and Continuous Colormap

While the optimal number of clusters for Washington, Texas, and California was $4$ (based on knee-point detection in BIC, see §5), we chose $5$ as the number of clusters for every state in our main analysis. Allowing for more clusters provides more granular information within urban areas while maintaining consistency with the $4$ cluster model. Figure S9 presents the clustering results with the optimal number of clusters for every state. Note that Figure 2 in the main manuscript provides more granular information for Washington, Texas, and California. Increasing the number of clusters beyond the optimal results in a finer partitioning of the embedding (Figure S10).

We also demonstrate that by modeling the data with a single dimensional parameter in the nonlinear embedding along the dense tubular manifold matches the intuition provided by increasing the number of clusters for the GMM. For example, we constructed a single dimensional phase variable along the manifold based on the cosine similarity of the data points in the 2D nonlinear embedding space. The result is an even smoother transition across urban, periurban, suburban, and rural areas consistent across all four states, see Figure S11.

### 7 Clustering in metropolitan areas: Georgia and California

Figures S12 and S13 present the clustering for metropolitan areas in Georgia and California, respectively.

### 8 Response Speed Distributions

We quantified the speed at which CBGs increased their stay-at-home behavior in response to the pandemic during a transition period between March and April (more specifically, March 10 – March 31) by the slope of a linear fit of the CBG mobility time series during the transition period

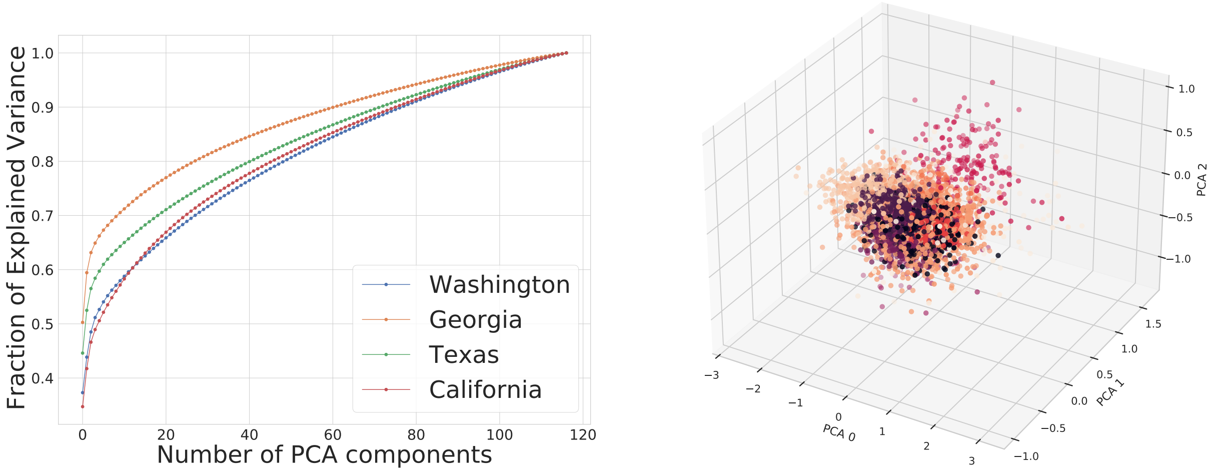


Figure S1: **Linear Dimensionality Reduction Performance.** Left: fraction of explained variance vs. number of PCA components for every state. Right: 3D visualization of PCA projection of the mobility time series data for Washington State with 11 clusters highlighted in color. Clustering was done in 8D PCA space.

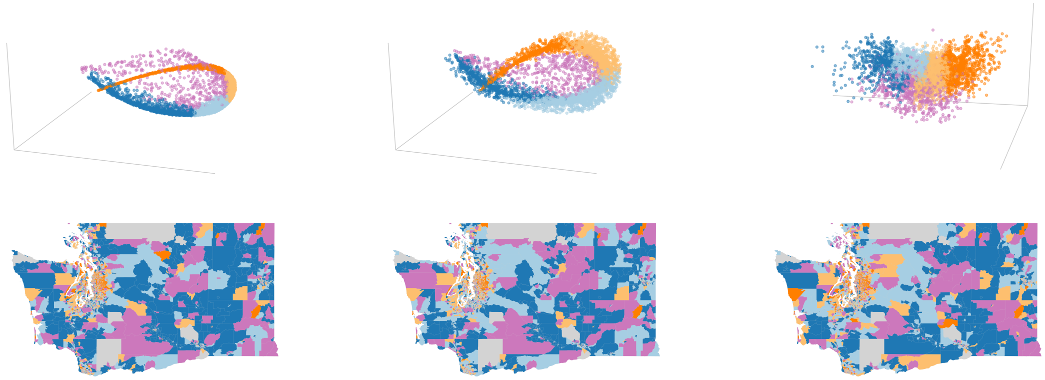

Figure S2: **Nonlinear Dimensionality Reduction Embeddings and Corresponding Maps for Washington State.** Top Left: Laplacian Eigenmaps, Top Middle: Locally Linear Embedding, Top Right: Isomap. Bottom Left: Laplacian Eigenmaps clustering map, Bottom Middle: Locally Linear Embedding clustering map, Bottom Right: Isomap clustering map.

(Figure S14). Figure S14 shows that the response speed distributions are directly correlated with CBG cluster assignment.

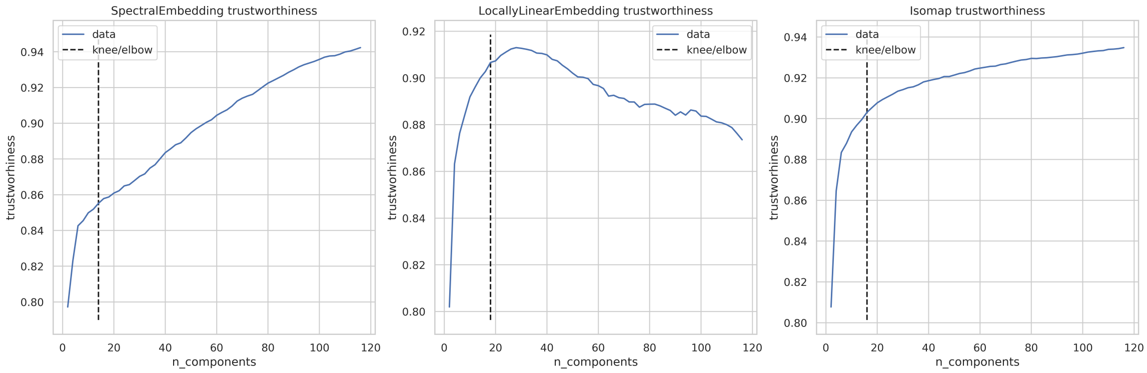


Figure S3: **Optimal Embedding Dimensionality for Washington State.** Left: Laplacian Eigenmaps (optimal dimensionality 14), Middle: Locally Linear Embedding (optimal dimensionality 12), Right: Isomap (optimal dimensionality 16).

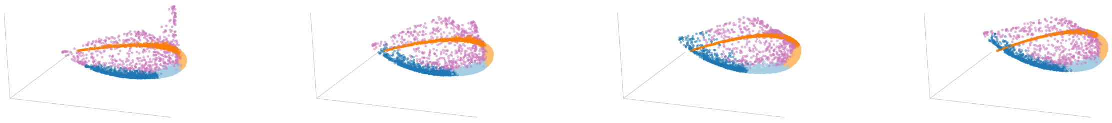

Figure S4: **Laplacian Eigenmaps Hyperparameter Robustness.** Each panel presents a 3D Laplacian Eigenmaps embedding of Washington state mobility data for `n_neighbors` in $\{20, 30, 40, 50\}$ respectively.

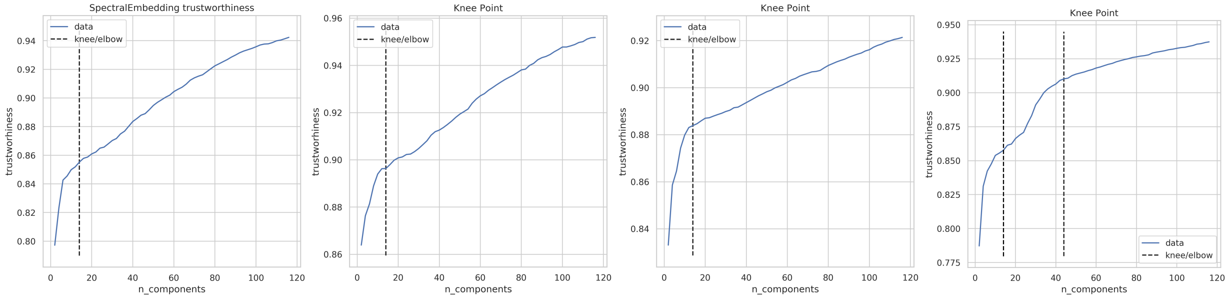


Figure S5: **Optimal Laplacian Eigenmap Embedding Dimensionality for Every State.** Each panel presents trustworthiness vs number of Laplacian Eigenmap components for Washington (optimal dimensionality 14), Georgia (optimal dimensionality 14), Texas (optimal dimensionality 14), and California (optimal dimensionality 14 or 44) respectively.

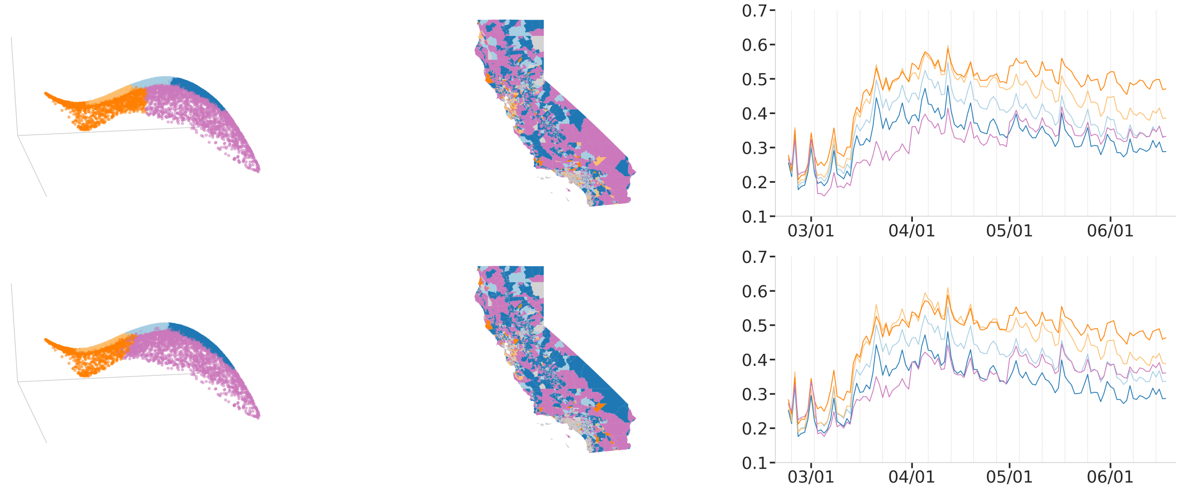


Figure S6: **Comparison of two trustworthiness knee points for California.** Top: Clustering results using 14D Laplacian Eigenmap embedding (3D illustration of the embedding, geographic map and average mobility time series per cluster with clusters highlighted in color). Bottom: Clustering results using 44D Laplacian Eigenmap embedding (3D illustration of the embedding, geographic map and average mobility time series per cluster with clusters highlighted in color).

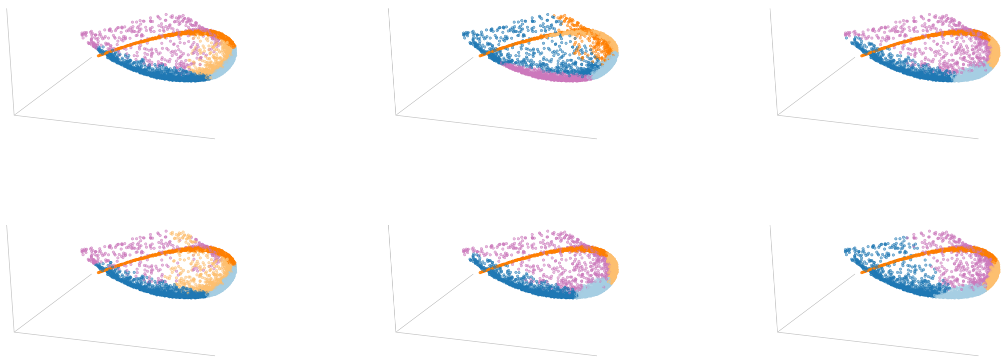

Figure S7: **Nonconvexity robustness.** Different types of GMM clustering results obtained by refitting GMM several times.

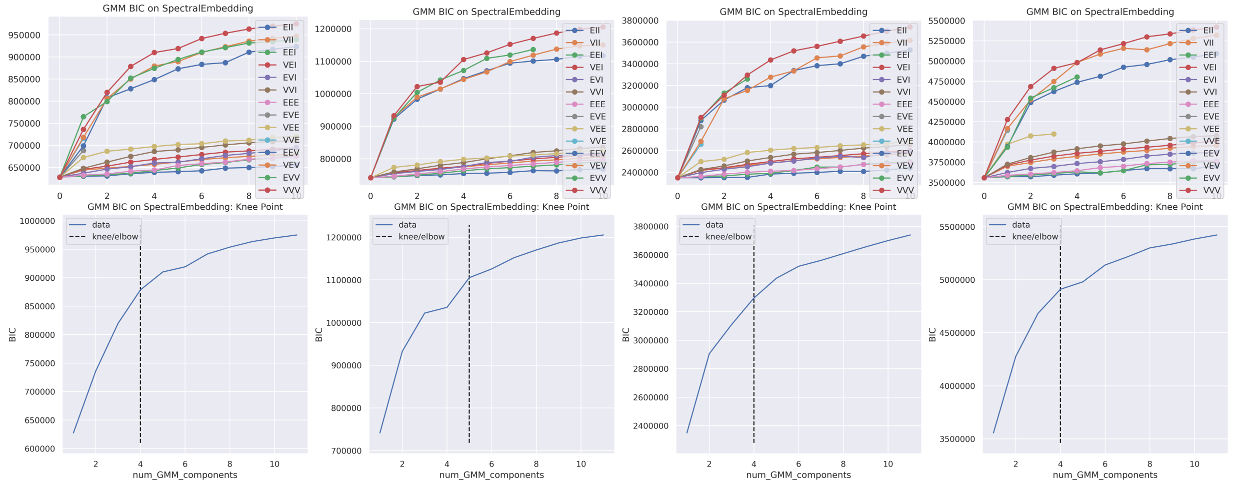


Figure S8: **GMM model selection for every state based on BIC.** Top: BIC curves for different parametrizations of the GMM model as described in [7]. Bottom: Optimal number of GMM components identified using knee-point detection on the best BIC curve for Washington (optimal number of clusters 4), Georgia (optimal number of clusters 5), Texas (optimal number of clusters 4), and California (optimal number of clusters 4) respectively.

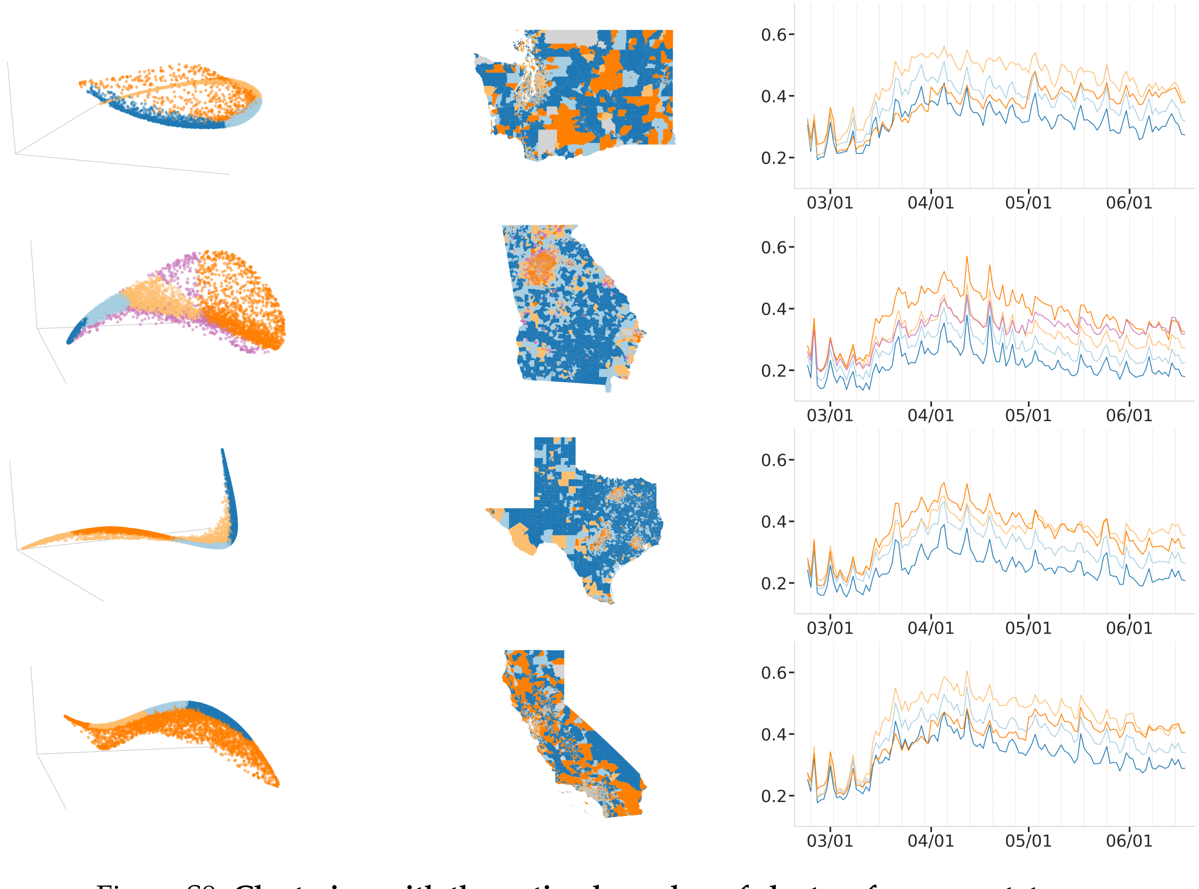


Figure S9: **Clustering with the optimal number of clusters for every state**

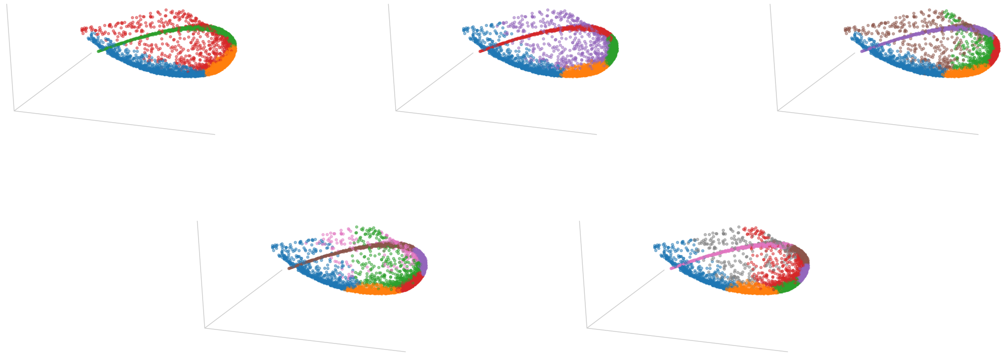

Figure S10: **Bigger number of clusters results in finer partitioning of the embedding.** Panels present clustering for the number of clusters in $\{4, 5, 6, 7, 8\}$ respectively.

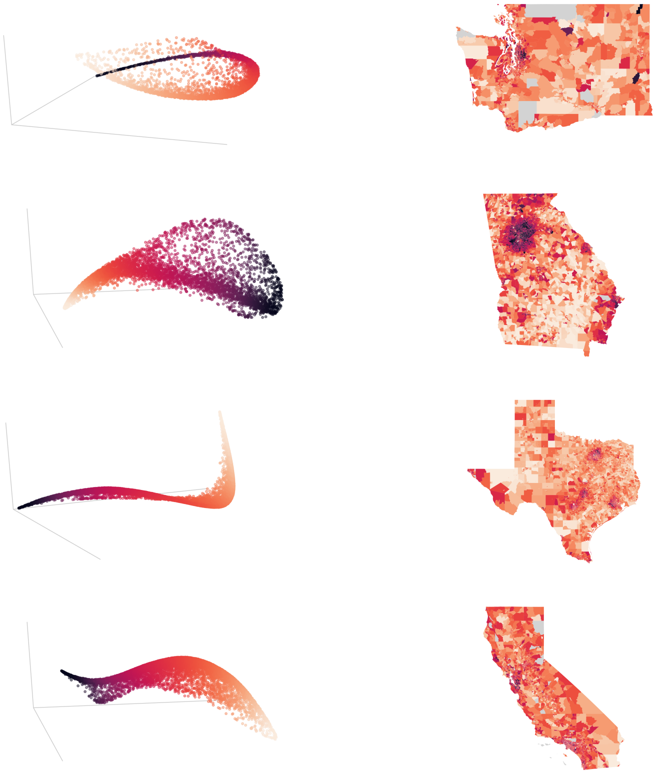

Figure S11: **Continuous Colormap.** Smooth transition across urban, periurban, suburban, and rural areas in Washington, Georgia, Texas, and California.

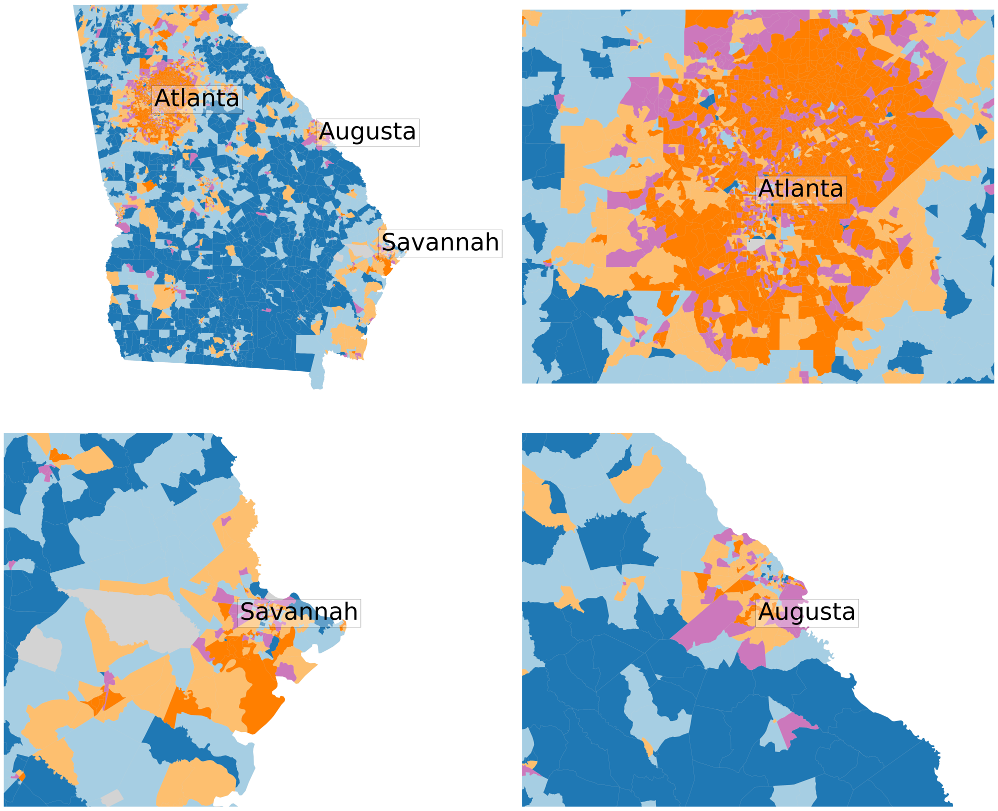


Figure S12: **Clustering in metropolitan areas in Georgia**

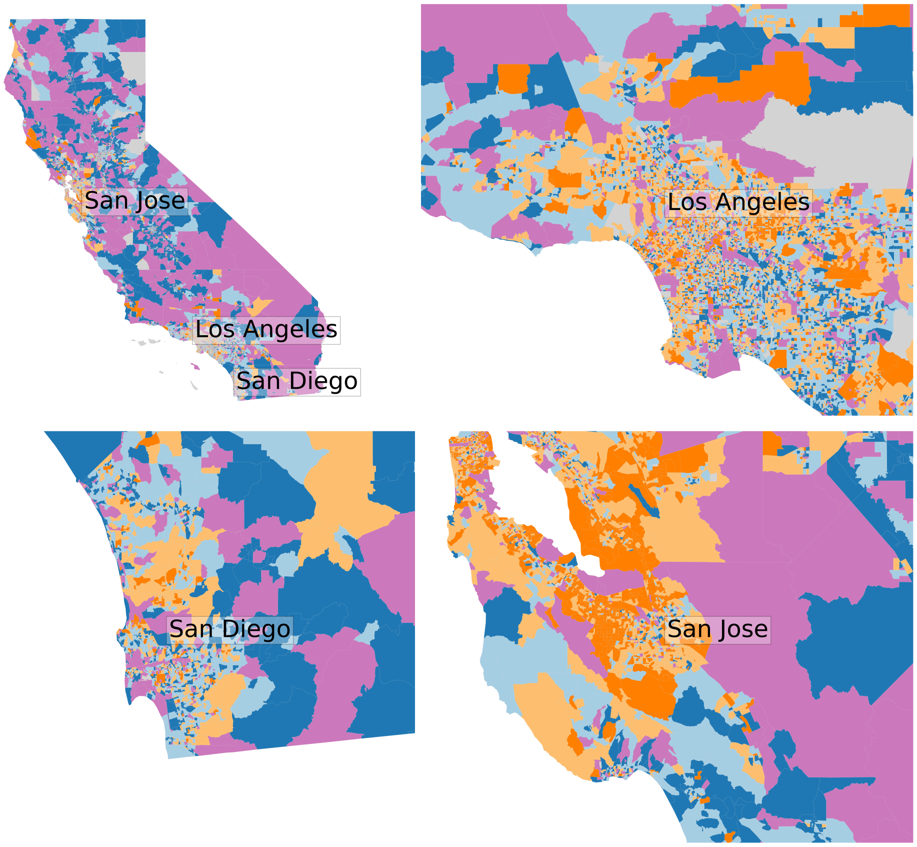


Figure S13: **Clustering in metropolitan areas in California**

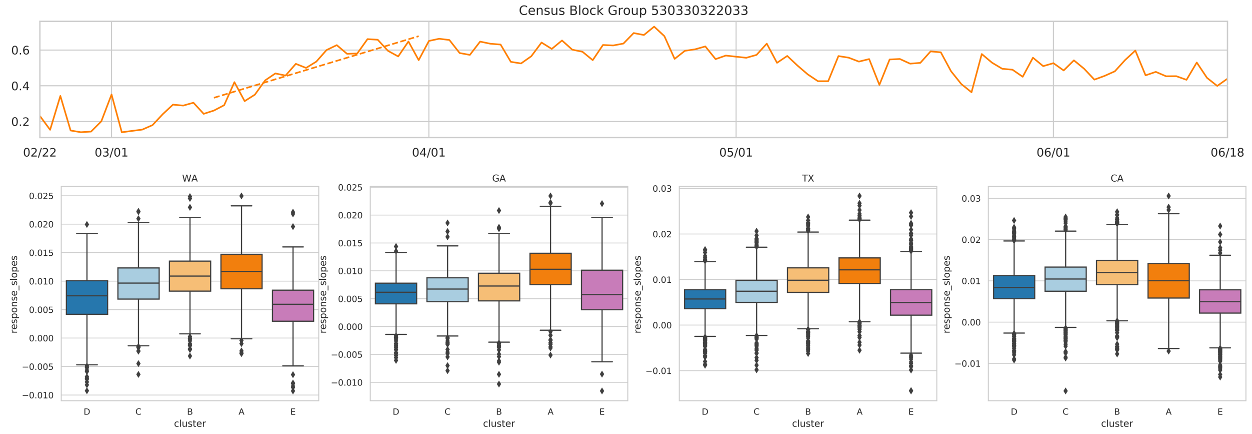


Figure S14: **Response Speed Distributions.** Top: The response speed is quantified by the slope of a linear fit of the CBG mobility time series during the transition period of March 10 – March 31, dashed line represents that linear fit for an example CBG. Bottom: Response speed distributions for every state.

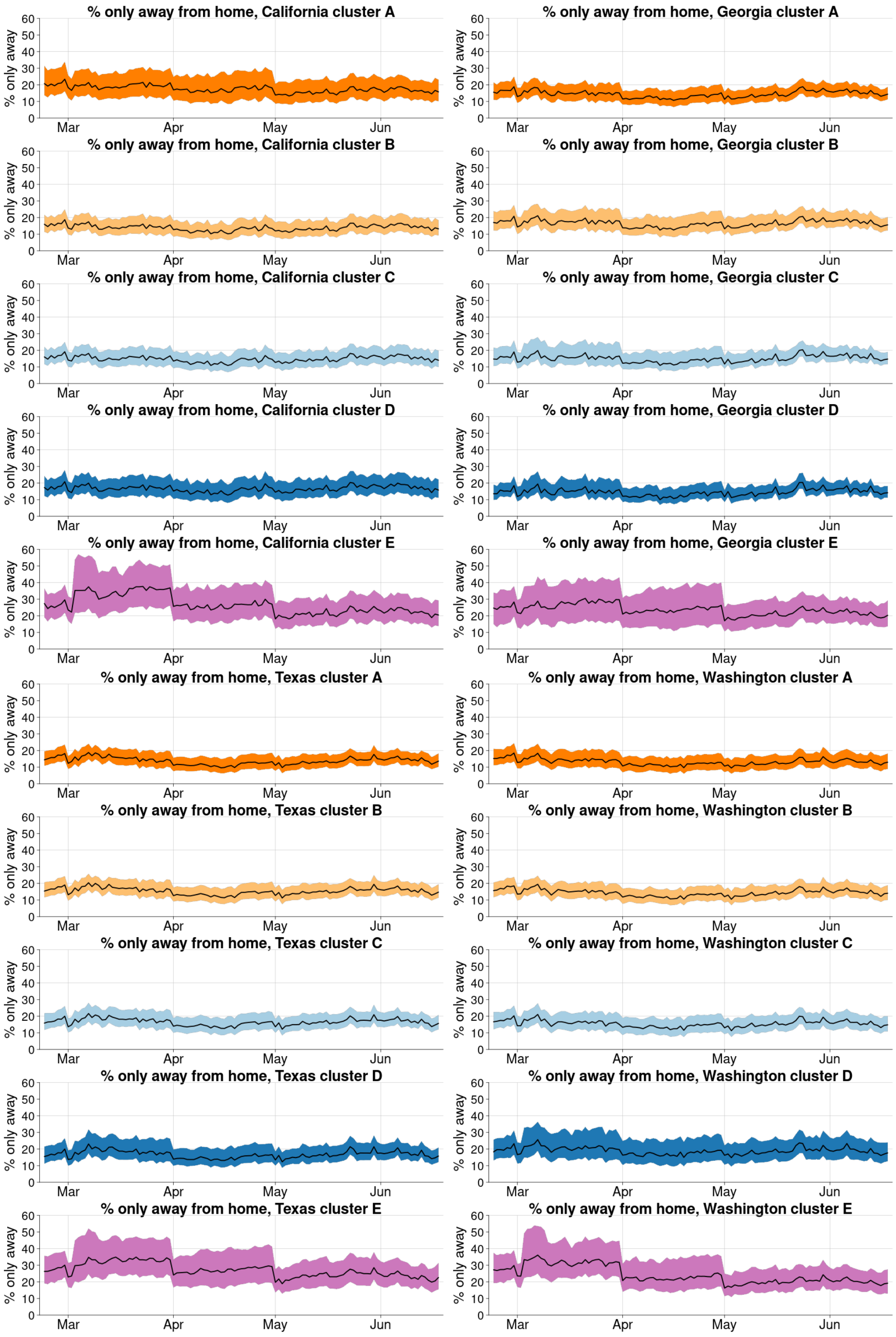


Figure S15: **The fraction of devices that are *only* away from their homes each day.** The medians and inter-quartile range are shown for each cluster.
